# Estimation of Rift Valley fever virus spillover to humans during the Mayotte 2018-2019 epidemic

**DOI:** 10.1101/2020.02.14.20022996

**Authors:** Raphaëlle Métras, W John Edmunds, Chouanibou Youssouffi, Laure Dommergues, Guillaume Fournié, Anton Camacho, Sebastian Funk, Eric Cardinale, Gilles Le Godais, Soihibou Combo, Laurent Filleul, Hassani Youssouf, Marion Subiros

**Author notes:** these authors share last authorship (Hassani Youssouf, Marion Subiros). Corresponding author: Raphaëlle Métras, Institut Pierre Louis d’Epidémiologie et de Santé Publique (iPLesp), Team 1: Communicable diseases Surveillance and Modelling, UMR-S 1136 | Inserm, Sorbonne Université, 27 rue de Chaligny 75012 Paris.

## Abstract

Rift Valley fever (RVF) is an emerging, zoonotic, arboviral haemorrhagic fever threatening livestock and humans mainly in Africa. RVF is of global concern, having expanded its geographical range over the last decades. The impact of control measures on epidemic dynamics using empirical data has not been assessed. Here, we combined seroprevalence livestock and human RVF case data from the 2018-2019 epidemic in Mayotte, with a dynamic mathematical model. Using a Bayesian inference framework, we estimated viral transmission potential amongst livestock, and spillover from livestock to humans, through both direct contact and vector-mediated routes. Model simulations were used to assess the impact of vaccination on reducing the human epidemic size. Reactive vaccination immunising 20% of the livestock population reduced the number of human cases by 30%. To achieve a similar impact, delaying the vaccination by one month required using 50% more vaccine doses, and vaccinating only humans required 20 times as more as the number of doses for livestock. Finally, with 53.92% (95%CrI [44.76-61.29]) of livestock estimated to be immune at the end of the epidemic wave, viral re-emergence in the next rainy season (2019-2020) was unlikely. We present the first mathematical model for RVF fitted to real-world data to estimate virus transmission parameters, and able to inform potential control programmes. Human and animal health surveillance, and timely livestock vaccination appear to be key in reducing disease risk in humans. We furthermore demonstrate the value of a One Health quantitative approach to surveillance and control of zoonotic infectious diseases.

## Introduction

Controlling zoonotic and vector-borne infections is complex, as it requires an accurate understanding of pathogen transmission within animal populations, and pathogen spillover to humans, whilst accounting for environmental factors affecting vector population dynamics (1,2). Rift Valley fever (RVF) is an emerging zoonotic arbovirosis causing haemorrhagic fever. RVF is a threat for both animal and human health, mainly in Africa (3). Livestock (cattle, sheep and goats) are RVF virus amplifying hosts, acquiring infection through the bites of infectious mosquitoes (mainly *Aedes* spp. and *Culex* spp.) (4). Humans get infected by direct contact with infectious animal tissues (upon abortions or animal slaughter), although vector transmission may also play a role (4,5). Since 2015, RVF has been listed as a priority emerging disease by the WHO R&D Blueprint (6). A major concern is the expansion of its geographical range over recent decades (5,7). Current disease control options for reducing disease risk in humans heavily rely on controlling virus transmission in animal populations. The impact of disease control measures in livestock on reducing RVF risk in humans has not yet been assessed, and doing so requires estimating key transmission parameters between livestock, and from livestock to humans; using animal and human epidemiological data.

Mayotte, an island located in the South Western Indian Ocean region, reported a RVF epidemic in 2007-2008 (8). In a previous paper, we used longitudinal livestock seroprevalence data to model RVF virus emergence in the livestock population, and we estimated that the likelihood of re-emergence was very low in a closed ecosystem (i.e. without introduction of infectious animals). However, a few imported infectious animals could trigger another large epidemic, as the herd immunity declined due to livestock population turnover (9). In 2018, about ten years after the previous epidemic, RVF outbreaks were reported in several East African countries (e.g. Kenya, South Sudan, Uganda, Rwanda) (10,11). In Mayotte, between November 2018 and August 2019, a total of 143 human cases (RVF virus RT-PCR confirmed) were reported (Fig. 1A). The virus belongs to the Kenya-2 clade (12), which is closely related to the strains detected in recent outbreaks in Eastern Africa. The Veterinary Services of Mayotte, the regional health authorities (Agence de Santé Océan Indien) and the French Public Health Agency (Santé Publique France) did further epidemiological investigations to assess temporal patterns in occurrence of the infection in the animal population, and to identify possible routes of human exposure to RVF virus. These investigations generated a uniquely well documented RVF epidemic dataset, including RVF seroprevalence and incidence data in animal and humans.

**Figure 1AF.**
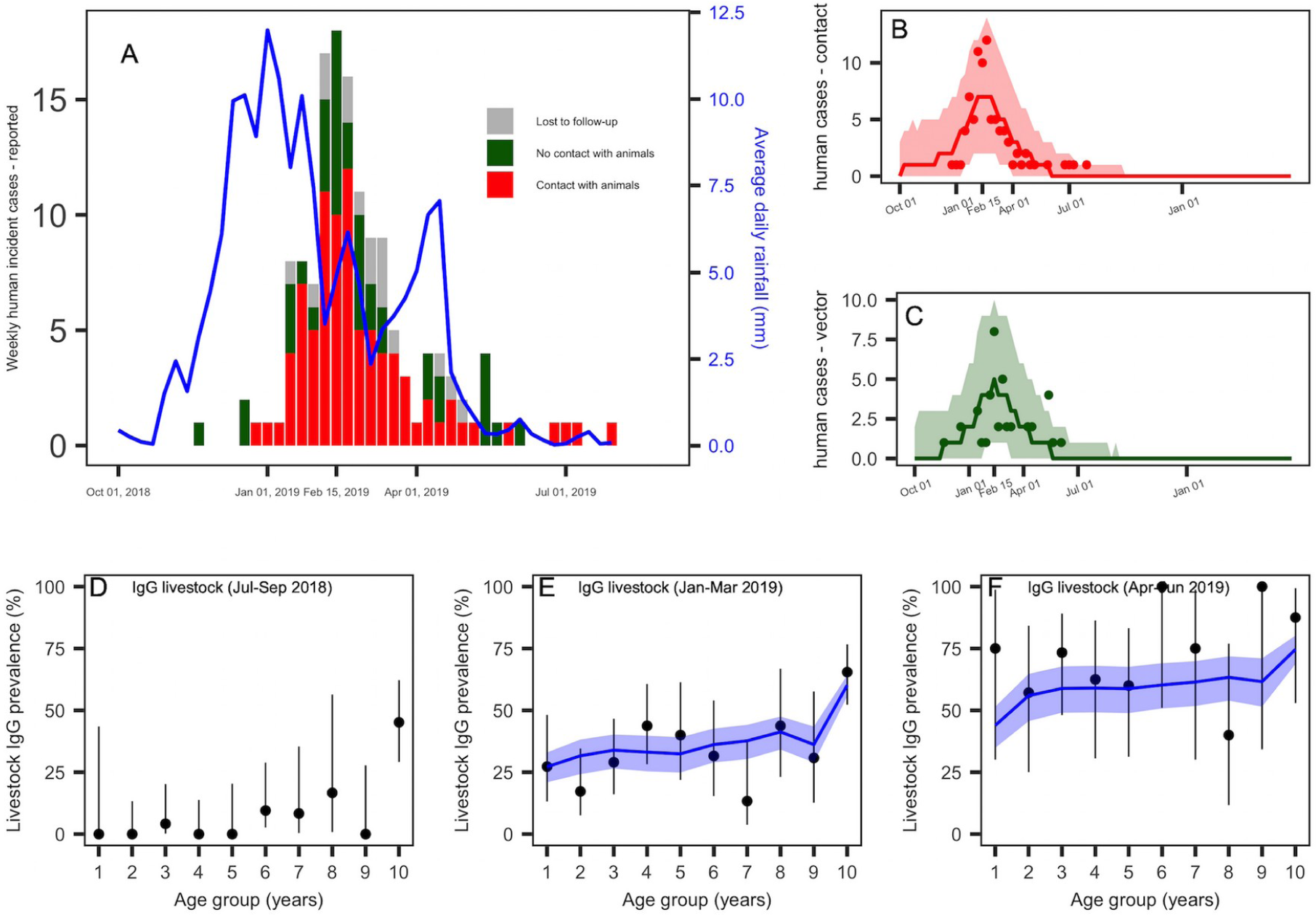
RVF epidemic data in humans and livestock, and model fit. *(A)* Weekly number of reported human cases and average daily rainfall pattern (solid blue line). Human cases reporting direct contact with animals or their products are presented in red (86 cases), those reporting no prior contact with animals or their products are in green (41 cases), and lost to follow-up are in grey (16 cases). *(B)* Predicted median (red solid line) and 95%CrI (red envelope) of the number of weekly reported human cases by direct contact, and weekly incident observed cases by contact (red dots). *(C)* Predicted median (green solid line) and 95%CrI (green envelope) of the number of weekly reported human cases by vector-mediated route, and weekly incident observed cases with no prior contact with animals (green dots). *(D)* Quarterly age-stratified RVF IgG seroprevalence in livestock for the trimesters July-September 2018 (N=173), *(E)* January- March 2019 (N=252), and *(F)* April-June 2019 (N=67). In *(D),(E),(F)*, the black dots and vertical black lines represent the observed age-stratified average IgG seroprevalence and their 95%CI. The model predicted values are showed by the median (solid blue line) and 95%CrI (blue envelopes).

We present these data and use them to extend and fit a mathematical model of RVF virus transmission in livestock (9), and explicitly account for viral spillover from livestock into the human population. We fit this model simultaneously to the infection patterns in livestock and human observed during the 2018-2019 epidemic, allowing for the first time, (i) to estimate the level of RVF virus transmission amongst livestock and spillover from livestock to humans by both direct contact and vector-mediated routes, (ii) to estimate the likelihood of another epidemic the following year, and (iii) to assess the impact of potential vaccination strategies in livestock and humans on reducing disease occurrence in humans.

## Results

### The course of the epidemic in livestock and humans

Between November 2018 and August 2019, 143 RVF human cases were reported. The epidemic peaked mid-February (February 11-17, 2019), with 18 weekly confirmed cases, six to seven weeks following the rainfall peak (Fig. 1A). About two-third of investigated cases reported a direct contact with livestock or its tissues (incl. milk consumption) (68%, n=86), whilst 32% (n= 41) reported no previous contact with animals (Fig 1A. cases in red and green, respectively).

Livestock sera (n=1,169) collected by the Veterinary Services between July 2018 and June 2019 were tested against RVF IgG. To assess the timing of emergence of the virus in the livestock population, we plotted quarterly age-stratified RVF IgG prevalence, using only tested animals for which the date of birth was available (n=493). In July - September 2018, that is before the report of the first human case, most seropositive animals were in the oldest age groups (Fig. 1D), possibly indicating viral exposure during the previous re-emergence (9). The IgG seroprevalence increased in all age groups in January-March (Fig. 1E), and then in April-June 2019 (Fig. 1F), evidencing that the emergence of the virus in the livestock population, was coincident with the report of cases in humans.

Ongoing viral phylogenetic analyses on human derived-samples (12), and IgM positive livestock seized from informal trade between June and August 2018 (Table S1), suggest that the virus was likely introduced from Eastern Africa into Mayotte between June and August 2018, through the movements of infectious animals.

### Epidemic model

We modelled virus transmission amongst livestock as a function of rainfall, therefore varying along the study period. RVF virus spillover from livestock to humans was modelled by both direct contact, assuming a time-invariant transmission rate, and vector-mediated transmission, defined as a function of rainfall ***(see Methods)***.

### Transmission parameters

By fitting this model to the RVF datasets, the time-varying reproduction number in the livestock population was estimated to peak at *R*_*s*_*(t)*=1.87 (95% Credible Interval CrI [1.53-2.69]), in the second half of January (January 14-27, 2019) (Fig. S3), two weeks following the rainfall peak, and three to four weeks prior to the predicted epidemic peaks in livestock and humans (Fig. 2A). This corresponded to a transmission rate amongst livestock (*β* _*L −L*_ (*t*)) at 8.92 per 100,000 livestock heads per day (95%CrI [1.09-7.77]). The spillover rate from livestock to humans by direct contact 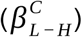 was estimated to 1.78 per 10 million persons per day (95%CrI [1.29 – 2.61]) (Table S2), and the maximum values of the time-varying spillover transmission rate 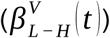 was 1.33 per 10 million persons per day (95%CrI [2.25-8.46]).

**Figure 2AC.**
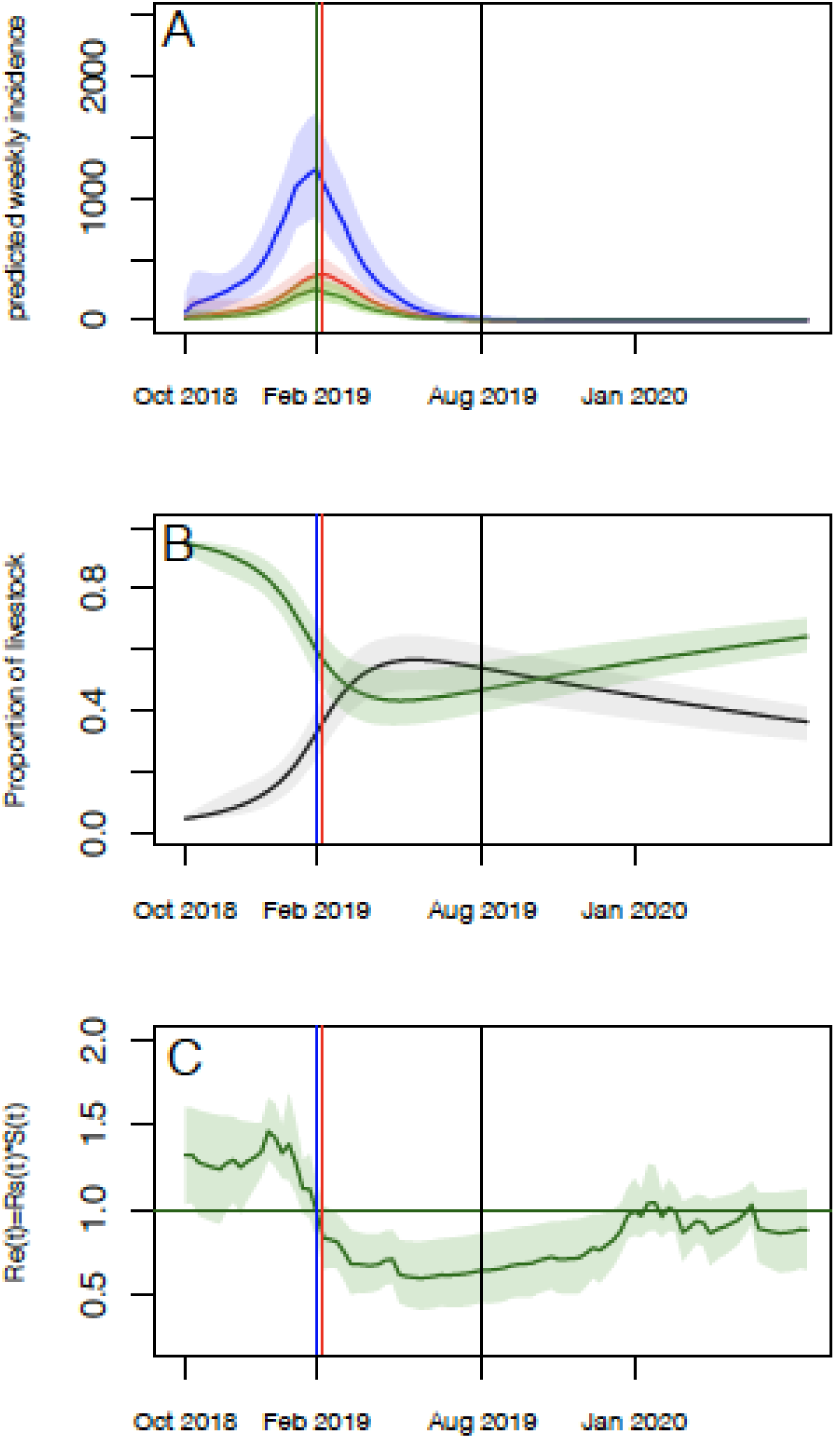
Model predictions over two rainy seasons (2018-2019 and 2019-2020): epidemic curves, proportion of susceptible and immune livestock, and time-varying effective reproduction (*R*_*e*_*(t)*) number. *(A)* Predicted (reported and unreported) number of infectious livestock (blue) and humans by direct contact (red), and vector-mediated route (green). *(B)* Predicted median (solid lines) and 95.%CrI envelopes of the predicted proportion of Susceptible (green) and Immune (black) livestock over the course of the epidemic. *(C)* Values of *R*_*e*_*(t)= R*_*s*_*(t)*S(t)* over the course of the epidemic. In all panels, the vertical blue and red vertical lines correspond to the predicted epidemic peaks in livestock and humans, respectively. The vertical black line corresponds to the end of the fitting period (August 2019).

### Model predictions

Using the estimated parameters, the simulated number of human reported cases was 181 (95% CrI [138-233]), with two third resulting from direct contact (n=111, 95%CrI [80-149]) and one third from vector transmission (n=70, 95%CrI [44-102]) (Table 1), in agreement with the observed data (Fig. 1B-1C). The predicted age-stratified IgG seroprevalence in livestock between January and June 2019 were in good agreement with the observed data as well (Fig. 1E-1F). The simulated incidence in livestock cases peaked mid-February (February, 11-17), concomitantly with the peak in human vector-mediated transmission, whilst the number of human cases by direct contact reached its maximum values one week later (February, 18-24) (Fig. 2A). Finally, by the end of the epidemic wave, 18,460 (95%CrI [14,926-21,154]) animals were affected resulting in 53.92 % (95%CrI [44.76-61.29]) of the livestock population being immune (Fig. 2B and Table 1). The overall predicted number of human cases (both reported and not reported) was estimated to have reached 9,566 (95%CrI [7,793-11,772]), resulting in 3.73% (95%CrI [3.03- 4.56]) of the human population being immune (Table 1).

**Table 1.** Predicted epidemic sizes (total and reported cases) and post-epidemic prevalence in humans and livestock, for the different scenarios, predicted median (95% Credible Interval, CrI).

| Scenarios | Epidemic size |  |  |  |  | Post-epidemic prevalence |  |
| --- | --- | --- | --- | --- | --- | --- | --- |
|  | Livestock total | Humans total | Humans reported | Reported contact | Reported vector | Livestock | Humans |
| 1. No intervention | 18,461<br>(14,926-21,153) | 9,566<br>(7,793-11,772) | 181<br>(138-233) | 111<br>(79-149) | 70<br>(44-102) | 53.92<br>(44.76-61.29) | 3.73<br>(3.03-4.56) |
| **Livestock vaccination** |  |  |  |  |  |  |  |
| 2. Dec 3,000 | 14,415<br>(11,154-17,237) | 7,465<br>(5,980-9,381) | 142<br>(106-186) | 87<br>(61-119) | 54<br>(34-80) | 42.40<br>(34.21-50.33) | 2.90<br>(2.33-3.65) |
| 3. Dec 6,000 | 11,447<br>(8,863-14,046) | 5,936<br>(4,683-7,674) | 113<br>(83-151) | 69<br>(46-97) | 43<br>(26-67) | 34.17<br>(27.66-41.15) | 2.31<br>(1.82-2.99) |
| 4. Jan, 3000 | 15,311<br>(12,237-17,985) | 7,867<br>(6,302-9,956) | 149<br>(112-197) | 91<br>(63-126) | 58<br>(35-84) | 44.72<br>(36.85-51.72) | 3.06<br>(2.45-3.88) |
| 5. Jan, 6000 | 13,229<br>(10,545-15,693) | 6,872<br>(5,470-8,752) | 131<br>(96-175) | 80<br>(55-112) | 51<br>(32-77) | 38.87<br>(32.21-45.44) | 2.68<br>(2.13-3.41) |
| 6. Jan, 9000 | 11,573<br>(9,296-13,784) | 6,014<br>(4,665-7,992) | 115<br>(83-158) | 90<br>(47-101) | 44<br>(27-68) | 34.27<br>(28.52-39.86) | 2.34<br>(1.82-3.12) |
| **Human vaccination** |  |  |  |  |  |  |  |
| 7. Dec, 50 % | 18,436<br>(14,987-21,099) | 6,032<br>(4,708-7,942) | 115<br>(82-157) | 70<br>(46-102) | 44<br>(26-67) | 53.79<br>(45.05-61.20) | 2.35<br>(1.83-3.10) |

In this setting, the likelihood of virus re-emergence the following rainy season (2019-2020) was less than 2.5% (Fig. 2A), with the time-varying effective reproductive number *R*_*e*_*(t)* falling below unity following the epidemic peaks and remaining very close to or below unity over the second year of the simulations (Fig. 2C).

### Vaccination scenarios

Probabilistic forecasts were also used to assess the impact of different livestock and humans vaccination strategies on the size of the epidemic in both animals and humans (Fig 3A-3D and Table 1). A reactive and mass vaccination campaign in livestock immediately after the report of the first human case (i.e. 6,000 doses in December 2018) allowed a reduction in the epidemic size by a third (median number of humans cases = 113 cases, median number of livestock cases = 11,447), while waiting one more month would have required 50 % more vaccine doses to achieve a similar impact (9,000 doses in January 2019, median number of humans cases = 115, median number of livestock cases = 11,573). Finally, a vaccination programme targeting only humans would require immunising half of the population (128,250 doses in December 2018) to reduce similarly the number of human cases (median=115 cases), whilst, of course, not impacting on the number of livestock cases.

**Figure 3AD.**
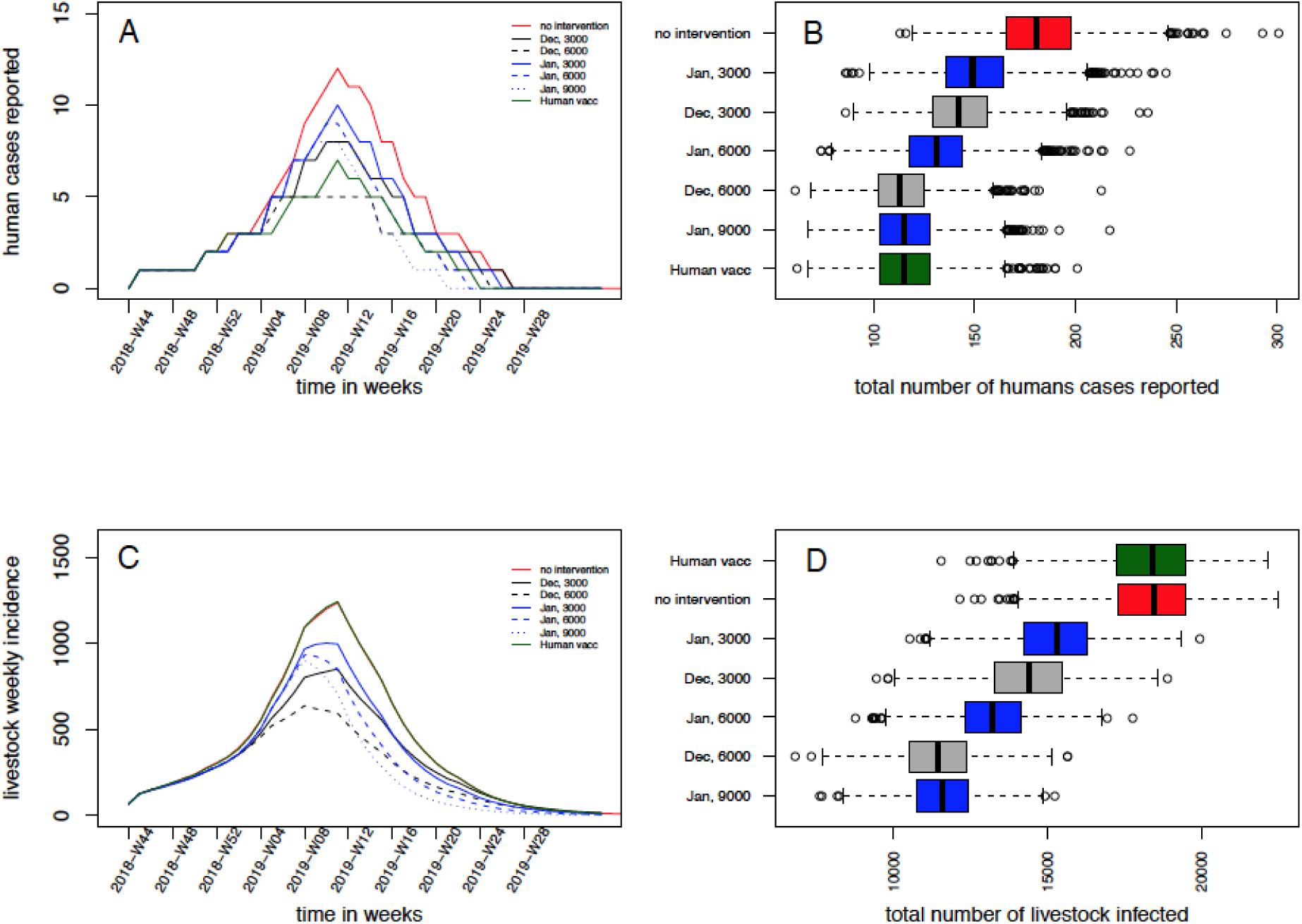
Impact of vaccination strategies on the epidemic size. *(A)* Median weekly number of predicted incident human cases, and corresponding *(B)* human epidemic size (reported cases). *(C)* Median weekly number of predicted incident infected livestock, and corresponding *(D)* total livestok epidemic size. In *(A)* and *(C)* the red solid line presents the scenario with no intervention (Scenario 1); the black lines present vaccinations in December 2018 (black solid: 3,000 doses, dashed black: 6,000 doses) (Scenarios 2-3); the blue lines present the vaccinations in January 2019 (blue solid: 3,000 doses, dashed blue: 6,000 doses; dotted blue: 9,000 doses) (Scenarios 4-6); the red line represent the human vaccination only (Scenario 7).

## Discussion

We present the first RVF epidemic dataset combining both livestock and human surveillance data, and use it to parameterise a mathematical model. We estimated, for the first time, transmission rates amongst livestock and spillover to humans using empirical epidemic data. This also allowed the quantitative assessment of the importance of timely livestock vaccination in reducing disease risk in humans during an epidemic, useful to inform potential control programmes, and illustrating the importance of One Health surveillance in the management of zoonotic diseases.

The IgM testing of illegally imported livestock suggested that the virus may have been introduced in Mayotte around June-August 2018, which is in agreement with the timing of RVF outbreaks on the East African mainland (11) and corresponds to the dry season in Mayotte. Viral transmission might have been maintained on the island at a low level in the dry season, or the virus might have been several times introduced, and the epidemic started only following the start of the rainy season (that is in October). The epidemic is likely to have therefore resulted from a recent viral re-introduction, rather that viral persistence over the last ten years, as concluded about the 2007- 2008 epidemics, in a previous study (9).

The systematic testing by RT-PCR of humans showing dengue-like syndrome performed in Mayotte for the last ten years (since 2008), provides additional evidence that RVF had been absent from the Island for a decade, and that the presented epidemic curve accurately reflected its actual timing. During the epidemic, mitigation strategies such as vector control around houses of human cases (i.e. post-detection) and the diffusion of prevention messages on milk consumption and exposure to animals were communicated, from February 27^th^ onwards (13), that is two weeks after the peak. Therefore, these measures are likely to have had a moderate impact on the epidemic size, whilst not affecting the time of the epidemic peak. In addition, the timing of the epidemic was corroborated by the observed changes in livestock seroprevalence, exhibiting a clear pattern of viral emergence. Most livestock sera (90%) were collected and tested as part of the regular annual surveillance campaign. As 10% of these samples were collected in areas reporting human cases, the proportion of seropositive animals may have been overestimated. However, most animal sampling was conducted from January 2019 onwards, when RVF virus had already spread across the whole island. In addition, our model predicted that 53.92 % (95%CrI [44.76-61.29]), of the livestock population was immune at the end of the simulated epidemic wave (August 2019), which was in line with estimates from the previous emergence in 2007-2008 (9).

Previous RVF models parameterised the transmission rate from livestock to humans by direct contact as an input parameter at 1.7 per 10 thousand persons per day (14-16). The epidemiological investigations conducted in this epidemic assessed whether human cases reported a direct contact with animals or their infectious tissues, and human cases without prior contact with such materials. This allowed for the first time estimating both RVF virus spillover to humans by direct contact and by vector transmission from epidemic data. These estimated transmission rates can be used as a benchmark for further modelling work. RVF human cases with or without previous contact with animals or animal products have been reported in other settings (17,18). Here, the reported fraction of cases without previous animal contact (32%) was three times higher than in South Africa (10%) (17). Several reasons may explain these differences. For example, this could result from a recall bias from people interviewed in Mayotte, or from the fact that in South Africa, people were tested following reports of RVF in animals (17).

Rainfall is a known driver for RVF virus transmission (19) and was used as a proxy for vector abundance. We assumed a 14-days lag between rainfall and its impact on vector abundance based on previous modelling studies on RVF vectors population dynamics (20,21). Temperature above 26°C may also promote RVF virus transmission (22-24). The temperatures of Mayotte varying annually between 25°C and 35°C (9), we assumed that in this specific setting, temperature would not be a major driver for viral transmission. In areas with cooler temperatures, such as South Africa (25), temperature may need to be taken into account (26). The highest estimated *R*_*s*_*(t)* value was 1.87 (95%CrI [1.53-2.69]), yet in line with previous estimations of *R*_*0*_ (14,27,28). The baseline model, with constant transmission parameters, had a similar DIC than the rainfall-dependent model, and showed *R*_*0*_ values within the same range. Whilst this may suggest a smaller influence of environmental factors on the RVF viral transmission dynamics during this epidemic, it may also suggest that upon the conditions met for emergence -(i) the presence of the virus, (ii) a susceptible livestock population and (iii) the presence of vectors - the epidemic fade-out likely resulted from a depletion of susceptible livestock. This was corroborated by the small likelihood of re-emergence in the following rainy season, with an effective reproductive number *R*_*e*_*(t)* remaining close or below unity in the months following the end of the 2018-2019 epidemic, due to the high proportion of immune animals.

A limitation of the model was that the reporting rate in humans was unknown, and defined based on data from the 2007-2008 epidemic (29). This relied on the assumption that both the 2007-2008 and 2018-2019 epidemics affected the same number of people. Whilst there is no data available on human infection patterns to support this assumption, our previous work estimated a post-epidemic livestock seroprevalence (9) which was similar to our current estimates, supporting the assumption that both epidemic sizes may be comparable. Further data collection estimating human post-epidemic seroprevalence would allow an accurate estimation of this reporting rate. Finally, the livestock model was built with similar assumptions than in our previous paper (9). This included a latent (E) and an infectious (I) period of 7 days in livestock, accounting for the extrinsic incubation period in the vector (3-7 days), and the latent (1-6 days) and infectious stages (3-6 days) in livestock (30-33), without explicitly modelling these processes. Although this may have slightly impacted on the predicted timing of the epidemic peak in humans, our model predictions were in agreement with the observations. In addition, this did not impact on the fitting with livestock data, as we fitted on the (R) compartment, aggregating data over three-month periods. We also assumed homogeneous mixing. Mayotte is a small island (374km^2^), the ecosystem shows limited spatial variations, livestock production systems are extensive with animals raised outdoor year round (9), compatible with the assumption that the livestock population was equally exposed to RVF mosquito vectors. Accounting for spatial heterogeneity, and testing for finer vaccination protocols would have required the use of epidemic data at a smaller spatio-temporal resolution. Our model can however be expanded into a metapopulation structure, and parameters further refined, in ecosystems with epidemic data available at finer spatial and temporal scales.

The impact and cost-effectiveness of livestock vaccination has been assessed in specific RVF high-risk areas in Kenya using simulation modelling (32,33). Instead, our analysis allows predicting the impact of vaccination strategies on reducing the number of human and animal cases, through a model calibrated from epidemic data. Our findings provide evidence that reactive animal vaccination is the most effective control measure, preventing both human and livestock cases, and requiring a smaller number of vaccine doses. The characteristics of the vaccine used in the vaccination scenarios (highly immunogenic, single dose, and safe) were those targeted by WHO R&D Blueprint (34), and not the existing ones. In practice, currently available RVF vaccines have different immunogenic and safety characteristics, with some of them requiring boosters (35), and the choice of which vaccine to use on the field may vary upon the epidemiological context. In addition, during this epidemic, livestock were not vaccinated due the absence of a vaccine with a EU marketing authorisation (Mayotte is an EU outermost region) (36). However, we highlight the importance of the development of contingency planning, availability of emergency funds and a suitable vaccine.

In conclusion, we have presented a uniquely detailed investigation into an outbreak of an emerging arbovirus, combining animal and human data, with a mathematical model for RVF. Early detection and rapid vaccination are critical to RVF control at the early stage of the epidemic. Disease surveillance in animals, contingency planning, and the timely implementation of livestock vaccination, are key for reducing human disease risk. This work represents a collaboration between public health agency, animal health surveillance network, farmers’ association, and researchers, initiated from the start of the epidemic, and conducted as a collaborative work as the epidemic unfolded. Delays in getting livestock data were inherent to climatic conditions (storms) and field work constraints in remote areas. Nevertheless, we addressed in practice the challenges of a quantitative One Health approach (37), and illustrated its value to surveillance and control of zoonotic emerging infectious diseases. Our model can be further expanded, refined and recalibrated for other ecosystems.

## Materials and Methods

### RVF datasets

#### Human data

Human incident case data were collected from patients showing dengue-like symptoms and consulting a GP, and who subsequently tested positive for RVF virus RT-PCR (38). Cases were interviewed using a structured questionnaire administered by Sante Publique France health epidemiologists (39). The number of incident cases was aggregated by week.

#### Livestock data

During the study period, livestock sera were sampled by field veterinarians according to two protocols: RVF targeted surveys around human cases and the regular annual surveillance campaign (SESAM) which is implemented since 2008 (8). The sera from the RVF targeted surveys were collected around human cases and were collected between January and March only. However, due to the rapidly increasing number of human cases and logistics constraints, the Veterinary Services instead requested field veterinarians to sample animals from the annual surveillance campaign only, depending on their regular field visits, therefore not depending on human cases. In total, between July 2018 and June 2019, a total of 1,169 livestock sera were collected (842 from the annual surveillance and 146 from human investigations), and tested against RVF IgG (ID Screen RVF Competition ELISA, IDVet, Grabels, France, Se=97 %, Sp=100 % (40)). Date of birth was available for 493 of these sampled animals (with 9% from RVF targeted surveys, Table S3). In order to follow the emergence of the virus in the livestock population over a year, we plotted quarterly age-stratified RVF IgG prevalence (Fig. 1D-1F).

#### Origin of the virus

To investigate the possible time window of virus introduction from imported infected animals, we collated serological data from illegally imported livestock seized by the Veterinary Services between June and August 2018. These animals were tested RVF IgM by ELISA (indicative of recent infections) (Table S1).

### Epidemic model

We modelled RVF epidemic from the start of the rainy season, the first week of October 2018 (October, 1-7), one month prior to the report of the first human case, up to the first week of August 2019 (July, 29-August, 4). No more human cases were reported after this date.

#### Transmission amongst livestoc

We adapted the previously developed SEIR model of RVF virus transmission amongst Mayotte livestock (9) to the current epidemiological context. For full details on the model structure and equations, see Metras *et al*. 2017 (9). We kept the previous underlying demographic livestock population age-structure (10 yearly age-groups) for fitting purpose, and we used a discrete-time deterministic framework, with a daily time step. In the previous model, the transmission parameter amongst livestock (*β* _*L −L*_ (*t*)) and corresponding time- varying reproductive number (*R*_*s*_*(t)*) were assumed to be vector-borne and modelled as a function of monthly NDVI (Normalized Difference Vegetation Index) values, as a proxy for vector abundance. Here, instead of using monthly NDVI, we used rainfall data (41) at a daily time step, since the model time step and the human epidemic curve available for fitting had a smaller time resolution. We also included a lag of 14 days between rainfall and its impact on the vector abundance (21,22). To look at the temporal pattern of the viral transmission over time, we also calculated *R*_*e*_*(t)*, the time-varying effective reproduction number, as the product of *R*_*s*_*(t)* with the proportion of susceptible livestock at time *t* (***SI Methods***).

#### Spillover into humans

We added a module simulating RVF virus transmission from livestock into the human population. We assumed that susceptible humans (*S*_*H*_) became infected following exposure with infectious livestock by direct contact 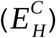 at a constant rate 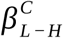, and by vector- mediated route 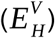 at a time-varying rate 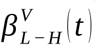, scaled on the rainfall-dependent within- livestock transmission (*β* _*L −L*_ (*t*)). Infected individuals 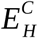 and 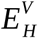 successively moved to their respective infectious states (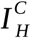 and 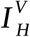); after which they moved into the immune compartment (*R*_*H*_) (Fig. S1), assuming they remained immune until the end of the study period. The model equations, transmission parameters and the formulation of the forces of infection from livestock *λ*_*contact*_*(t)* and vectors *λ*_*vector*_*(t)* are presented in ***SI Methods***.

#### Parameterisation and model fitting

Input parameters were those related to the natural history of infection and demographics in both livestock and human populations (Table S4). The proportion of immune animals at *t*_*0*_ was informed from the aggregated July-September 2018 IgG livestock seroprevalence campaign (Fig. 1D). The reporting fraction of human cases was set to *ρ*=1.9%, as a post-epidemic serological study in humans, conducted in 2011 in Mayotte, estimated that 3.5% (95%CI [2.6-4.8]) of the human population was RVF IgG-positive (30). Assuming a population size of 212,645 inhabitants in 2012 (42), this corresponded to an average of 7,442 persons being seropositive. Assuming that the sizes of the 2007-2008 and 2018-2019 epidemics were similar, the detection of 143 cases in the 2018-2019 epidemics suggests a reporting fraction of 1.9% (95% CI [1.4-2.6]). Finally, input rainfall data were downloaded from the Meteofrance website, as cumulated rainfall over 10-day periods (41). Daily rainfall was calculated by dividing these values by ten over each 10-day period.

Five parameters were estimated by fitting the model to the human and livestock epidemic data (Table S3). Two parameters related to the rainfall-dependent transmission amongst livestock (*A* and *B*), two parameters estimated the spillover to humans, via contact with livestock 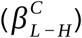 and via vector (scaling factor *X*), and the fifth parameter was the number of infectious livestock at *t*_*0*_ (*I*_*liv0*_). Parameter estimation was done by fitting simultaneously the (i) quarterly age-stratified simulated proportion of immune livestock (*p*_*a,q*_) to quarterly RVF IgG prevalence (Fig. 1E-1F); (ii) the simulated weekly number of reported incident cases in humans by direct contact (Fig. 1B) and (iii) the simulated weekly number of reported incident cases in humans by vector-mediated route (Fig. 1C), to the observed weekly number of reported cases via both transmission routes. Values of those five parameters were sampled from their prior distribution 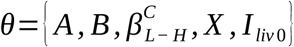 using a Monte Carlo Markov Chain Metropolis-Hastings (MCMC-MH) algorithm, implemented in the fitR package (43). Finally, to assess the impact of rainfall over the course of the epidemic, we also fitted a baseline model for which all transmission parameters were constant over time (Table S5). Details on models equations, parameter estimation, model fitting, and model comparison are presented in ***SI Methods***.

#### Forecasting and vaccination scenarios

We did probabilistic projections for seven scenarios (Table 1). For all scenarios, we simulated 2,500 stochastic trajectories by sampling randomly parameter values from the joint posterior distribution. Scenario 1 aimed at estimating the likelihood of virus re-emergence, without disease control intervention, in the following rainy season (in 2019-2020), in a closed ecosystem, using the same rainfall data as during the 2018- 2019 rainy season. Scenarios 2-6 aimed at assessing the impact that different livestock vaccination strategies could have had on the number of human and livestock cases during the 2018-2019 epidemic. We assumed the use of a single-dose highly immunogenic vaccine (90 % vaccine efficacy) (34,35), and a 14-days lag between vaccination and build-up of immunity. Figures of vaccination campaigns in Mayotte in 2017 (against blackleg, a livestock disease), showed that about 3,000 vaccine doses are routinely administered to livestock over a year by local veterinarians. Scenario 2 tested the impact of administrating all these 3,000 doses in one month, in December 2018, immediately after the report of the first human case (joint animal-human alert date for response), corresponding to the current vaccinating capacity in Mayotte in an emergency setting. Scenario 3 assumed an extra-vaccine supply and an emergency mass vaccination, allowing 6,000 doses to be administered in December 2018. We also assessed the impact of vaccinating livestock in January 2019, one month following the report of first human case, allowing extra time for organising the vaccination campaign : 3,000 doses (Scenario 4), 6,000 doses (Scenario 5) and 9,000 doses (Scenario 6). Finally, to assess the impact of a reactive and mass vaccination only in humans, we simulated a 50% vaccination coverage of the human population in December 2018 (i.e. 128,250 doses) (Scenario 7). Vaccination equations and diagram are presented in ***SI Methods*** and Fig. S2.

## Data Availability

Aggregated data will be made available by the authors following publication in a peer-review journal

## Acknowledgments

The authors wish to thank the Agence de Santé océan Indien that has participated in collecting human cases data, the laboratory of Centre Hospitalier de Mayotte which has performed the virological analyses on human samples, the animal SESAM (Système d’Epidémiosurveillance Animale à Mayotte) surveillance system, the CoopADEM (Coopérative agricole des éleveurs mahorais), the Cirad-CYROI, the Veterinary Services, and the LVAD (Laboratoire Vétérinaire d’Analyses Départemental de Mayotte) for the data collection and the serological analyses on livestock samples. Finally, we thank Harold Noël from Santé publique France for facilitating human data access in the early stage of the epidemic.

## Author Contributions

RM, WJE, LD, YH, MS conceptualized and designed the study. CY, LD, SC, YH, MS collated the data and did data management. RM, GF performed the analyses. RM, WJE, CY, LD, GF, AC, SF, GLG, CS, EC, LF, YH, MS interpreted and discussed the data and results. RM was responsible for drafting the manuscript. All authors reviewed and approved the final manuscript.

## Ethics statement

The livestock data were collected under the under the Mayotte disease surveillance system (Système d’Epidémiosurveillance Animale à Mayotte, SESAM) with the approval of the Direction of Agriculture, Food and Forestry (DAAF) of Mayotte. For human data, according to French law, only “research involving a human being” (research defined by article L. 1121–1 and article R. 1121–1 of the Code de la santé publique) are compelled to receive the approval of ethics committee. This study was based on anonymous data collected from health professionals for public health purposes relating to the health surveillance mission entrusted to Santé publique France by the French Law (article L. 1413-1 code de la santé publique). Therefore, the study did not meet the criteria for qualifying a study “research involving a human being” and did not require the approval of an ethics committee. Furthermore, as the data were anonymous, it did not require an authorization of the French data protection authority (Commission Nationale informatique et libertés).

